# Transcriptome landscape of high and low responders to an inactivated COVID-19 vaccine after 4 months using single-cell sequencing

**DOI:** 10.1101/2024.04.07.24305443

**Authors:** Zhongyi Zhu, Yaling Huang, Jiatong Sun, Meirong Li, Yong Chen, Lei Zhang, Fubaoqian Huang, Chuanyu Liu, Weijun Chen, Jinmin Ma

## Abstract

**Background:** Variability in antibody responses among individuals following vaccination is a universal phenomenon. Single-cell transcriptomics offers a potential avenue to understand the underlying mechanisms of these variations and improve our ability to evaluate and predict vaccine effectiveness.

**Objective:** This study aimed to explore the potential of single-cell transcriptomic data in understanding the variability of antibody responses post-vaccination and its correlation with transcriptomic changes.

**Methods:** Blood samples were collected from 124 individuals on day 21 post COVID-19 vaccination. These samples were categorized based on antibody titers (high, medium, low). On day 135, PBMCs from 27 donors underwent single-cell RNA sequencing to depict the transcriptome atlas.

**Results:** Differentially expressed genes (DEGs) affecting antibody expression in various cell types were identified. We found that innate immunity, B cell, and T cell population each had a small set of common DEGs (MT-CO1, HLA-DQA2, FOSB, TXNIP, and JUN), and Macrophages and Th1 cells exhibited the largest number of DEGs. Pathway analysis highlighted the dominant role of the innate immune cell population in antibody differences among populations, with a significant impact from the interferon pathway. Furthermore, protein complexes analysis revealed that alterations in the ribosome complex, primarily regulated by DC cells, may play a crucial role in regulating antibody differences. Combining these findings with previous research we proposed a potential regulatory mechanism model of DC cells on B cell antibody production.

**Conclusion:** While direct prediction of specific antibody levels using single-cell transcriptomic data remains technically and data-wise challenging, our study demonstrated the vast potential of single-cell transcriptomics in understanding the mechanisms underlying antibody responses induced by vaccines.

## Introduction

Vaccine efficacy is often gauged by measuring the levels of specific antibodies generated post-vaccination, which play a crucial role in identifying and neutralizing pathogens, thus preventing diseases. Studies have consistently demonstrated a positive link between vaccine-induced antibody levels and their protective capacity, highlighting the importance of post-vaccination antibody assessment[1–4]. Therefore, measuring the antibody levels after vaccination has become a key indicator for assessing vaccine effectiveness. However, it’s common to observe variations in antibody levels within the population following vaccination. While most individuals exhibit an “average” antibody response, a subset of individuals generates exceptionally high antibody levels (high-responder), while others display low antibody levels (non-responder). This phenomenon, attributed not solely to the type of antigen of a vaccine, has also been observed across successful COVID-19 vaccines, including mRNA and inactivated vaccines[5–8].

Numerous studies have investigated the variation of antibody responses in populations following vaccination. It is generally believed that these differences can be attributed to two main factors. Firstly, vaccine-related factors such as antigen type, vaccine formulation, type of vaccination[9], number of doses, and vaccination schedule[10] play a role. Secondly, host-related factors including age, gender, weight, health status[11], and genetic background differences are also influential[5, 12]. In terms of research focus, there is a greater emphasis on optimizing vaccination programs than on exploring the mechanisms of host variations. This may be partly due to the statistical challenge of eliminating the interference of various vaccine formulations on inter-host differences. However, for COVID-19 vaccines, studies have already demonstrated the impact of host factors such as age[13], medical history[14], and genetic background[15, 16] on post-vaccination antibody levels. Nevertheless, these studies mainly report statistical aspects, with limited exploration of underlying mechanisms.

Generally, the production of specific antibodies in peripheral blood after vaccination is a complex process involving the activation of the immune system. In this process, innate immune cells first recognize the antigen, followed by the activation of B cells, which then differentiate into plasma cells in the peripheral blood and produce a large number of antibodies targeting specific pathogens[17, 18]. However, the activation of B cells is not a single step but rather a process involving multiple cells and complex transcriptional regulation interactions. Single-cell transcriptomic sequencing technology provides a powerful tool for in-depth understanding of transcriptional interactions between cells in peripheral blood[19, 20]. Through detailed and comprehensive transcriptomic analysis of peripheral blood mononuclear cells (PBMCs), this technology can reveal the dynamic changes in cell heterogeneity and their transcriptional regulation under different health conditions. Therefore, analyzing single-cell transcriptomic data from different response groups after vaccine administration can help to detail the key features influencing vaccine antibody responses at the peripheral blood transcriptomic level. However, in practical applications, single-cell transcriptomic sequencing technology still faces several challenges[21, 22]. One of these challenges is establishing a unified method for classifying and annotating cell types across different organs and tissues. Even in the single-cell transcriptomic relatively well-studied field PBMCs, previous single-cell transcriptomic studies have often shown inconsistency in cell type annotation, and the use of marker sets lacks uniform standards. While efforts are currently underway in the research to develop standardized reference databases and bioinformatics-based unified annotation methods, these approaches are still in the developmental stage and have yet to be widely implemented and adopted[23, 24].

The aim of this study is to investigate the immunological mechanisms underlying the differences in antibody responses by analyzing the transcriptional characteristics of different antibody response groups four months after receiving the inactivated COVID-19 vaccine. To accurately identify cell types, we employed a traditional flow cytometry-based approach, focusing primarily on identifying known peripheral blood mononuclear cell (PBMC) populations rather than discovering new cell subtypes. Our goal is to analyze the transcriptomic differences among classical cell subtypes and reveal key cell types, transcriptional regulatory factors, and immune regulatory networks that may influence antibody responses. This will provide deeper insights into understanding and supporting immune protection following vaccination. Additionally, we have developed a flow cytometry-based single-cell annotation method that has shown good application performance in this study, and we anticipate its widespread use in future research.

## Results

### Result 1: Identifying PBMC Cell Subtype Profiles of Antibody Response Groups by Reference to Flow Cytometry Markers

Sine the purpose of this study was to characterize the transcriptomic profile of different antibody response groups at the single-cell level. We divided the antibody response population into high, medium, and low groups based on a normal distribution of antibody data from 124 healthy volunteers. These volunteers had received two doses of the COVID-19 inactivated vaccine and were tested for antibody titers on day 21 after the second dose (Fig1A). We defined the middle response group as comprising 80% of the population (Fig1H), and then selected a total of 27 samples (4 from the high-response group, 19 from the middle-response group, and 4 from the low-response group) to collected blood samples at 4 months for antibody testing, PBMC isolation and transcriptome sequencing. We also confirmed that the statistical differences in antibody response were consistent at Day 21 and Day 135, and these differences were independent of age and sex composition. (Fig1I and Table S1).

Subsequently, we aimed to tackle the challenge of annotating cell types in PBMC single-cell transcriptome data. As outlined in the background section, the markers utilized in different studies are generally inconsistent, leading to variations in annotated cell types. To address this issue, we first compiled a list of classic cell type markers commonly used in flow cytometry (Table S2) and then explored the annotation effects of different marker combinations. Furthermore, in line with the objectives outlined in the background, our study specifically focused on accurately identifying classical PBMC cell types for improved analysis and comparison with previous flow cytometry-based results. This thought is particularly relevant as flow cytometry analysis remains the primary method for identifying cell types and investigating cell type-based mechanisms, including cytokine expression in vaccine studies[25, 26]. Hence, our study did not delve into the exploration of potential new cell subtypes in peripheral blood PBMC, as our primary focus was on the identification of classical PBMC cell types.

It might have been easy to anticipate, but our exploratory analysis revealed the challenge of accurately annotating cell subtypes using a single set of markers, particularly for classical B cell and T cell subtypes. The characteristics of positively identified cells in annotated clusters often do not align fully with theoretical flow cytometry identifications. This discrepancy can be attributed to differences between transcript expression and protein expression, as demonstrated by CD45RA and CD45RO. In these cases, gene expression typically only detects CD45 (PTPRC) gene expression at the mRNA level, making it challenging to directly distinguish between the two protein isoforms. While some solutions have been explored for this issue[27], the overall usability remains challenging and requires further improvement.

Finally, we utilized 4 sets of markers for primary and subcellular cell type annotation (Table S2). By employing a method that annotates small group subdivisions from large group annotations, we annotated the PBMCs into 21 cell subtypes in this study (Fig1B-F). Detailed illustration can be found in the Methods section (FigS1-S7). Our annotation includes 5 types of innate immune cells, including NK cells, macrophages, monocytes and DC cells (FigS8-S9). 4 B cell subgroups, including naive, plasma, memory and B regulatory cells (FigS10-S11), and 13 T cell subgroups (CD4, CD8, naive, memory, regulatory T cells, helper T cells, cytotoxic T cells) (Fig S12-S15). We compared the proportions of these cell types among the high, medium, and low antibody titer groups and found most cell type showed no significant differences in their overall distribution (Figure 1G). However, plasma cells with high expression of IL-6 showed varying proportions between the normal and high antibody titer groups (Figure S12). Considering that the samples were collected four months after vaccination, we expected the major cell types to be in a relatively stable state. However, minor differences may still exist in the proportions of certain cell subtypes. Overall, our analysis results align with this expectation.

**Figure 1:**
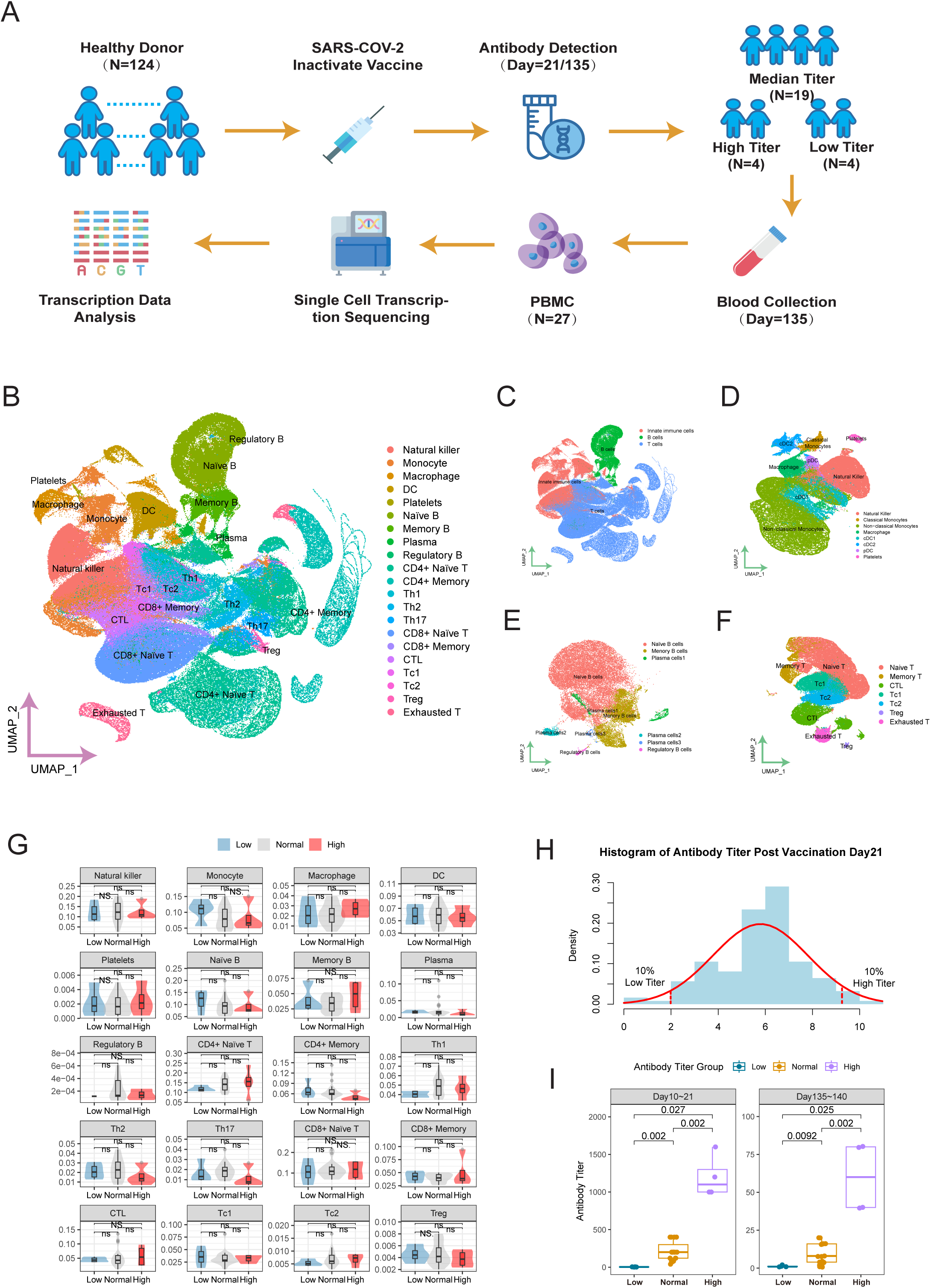
Transcriptomic Profiles of Individuals with Different Antibody Titers Four Months Post SARS-CoV-2 Vaccination. (A) Overview of Study Design. A total of 27 participants were enrolled in this study. They were classified into high, median, and low groups based on the antibody titers measured on day 21 and day 135 post-vaccination. All samples’ PBMCs on day 135 were subjected to Single-Cell Transcriptomics Sequencing. (B-F) Overall Transcriptomic Landscape of PBMCs from 27 Samples. An integrated transcription map from all participants (B), transcription map colored by three cell types (C), Innate Immune Cells (D), B cells (E), and CD8+ T Cells (F). (G) Cell Proportion Analysis Across 27 Cell Clusters. The x-axis represents the sample grouping, and the y-axis represents the cell proportion. (H) A histogram displaying the antibody titers on Day 21 post-vaccination, based on data from 124 healthy donors who received two doses of the COVID-19 inactivated vaccine. (I) Graph illustrating the differences in antibody titers between groups, with group categories on the x-axis and antibody titers on the y-axis. Each panel represents the time interval after the second vaccine dose. The differences between each pair of groups are calculated using Wilcoxon test and displayed as p-values.

In general, the proportions of various cell subtypes were consistent with our expectations (Figure 1G). Additionally, we specifically focused on the proportions and ratio of Th1 and Th2 cell subtypes. The evaluation of Th1 and Th2 cell proportions is commonly used to assess vaccines, particularly for infectious diseases like COVID-19 and RSV, where the Th1/Th2 ratio is indicative of the likelihood of vaccine-enhanced respiratory disease (VERD). Our results showed that in the evaluated samples, the proportion of Th1 cells was 0.27% of total PBMCs and 6.78% of CD4+ T cells, while the proportion of Th2 cells was relatively low (0.13% of total PBMCs and 3.26% of CD4+ T cells). This suggests a prevailing Th1 bias in the cellular antibody response four months after the administration of the COVID-19 booster vaccine. These findings indicate the potential of single-cell transcriptomic analysis as a complementary method for assessing cellular antibody responses following vaccination.

### Result 2: Macrophages and Th1 cells exhibit the highest number of genes contributing to antibody variances

For each cell type we identified, differential gene analysis was conducted by comparing two groups (both low vs median and median vs high groups). Upon aggregating all identified differential genes for analysis, we found that cell subtypes in PBMC with the most genes affecting antibody titer after vaccination were macrophages (112 genes), Th1 cells (104 genes), memory B cells (75 genes), as well as two types of DC cell subtypes (cDC, 68 genes; and pDC, 65 genes) (Fig2F).

**Figure 2:**
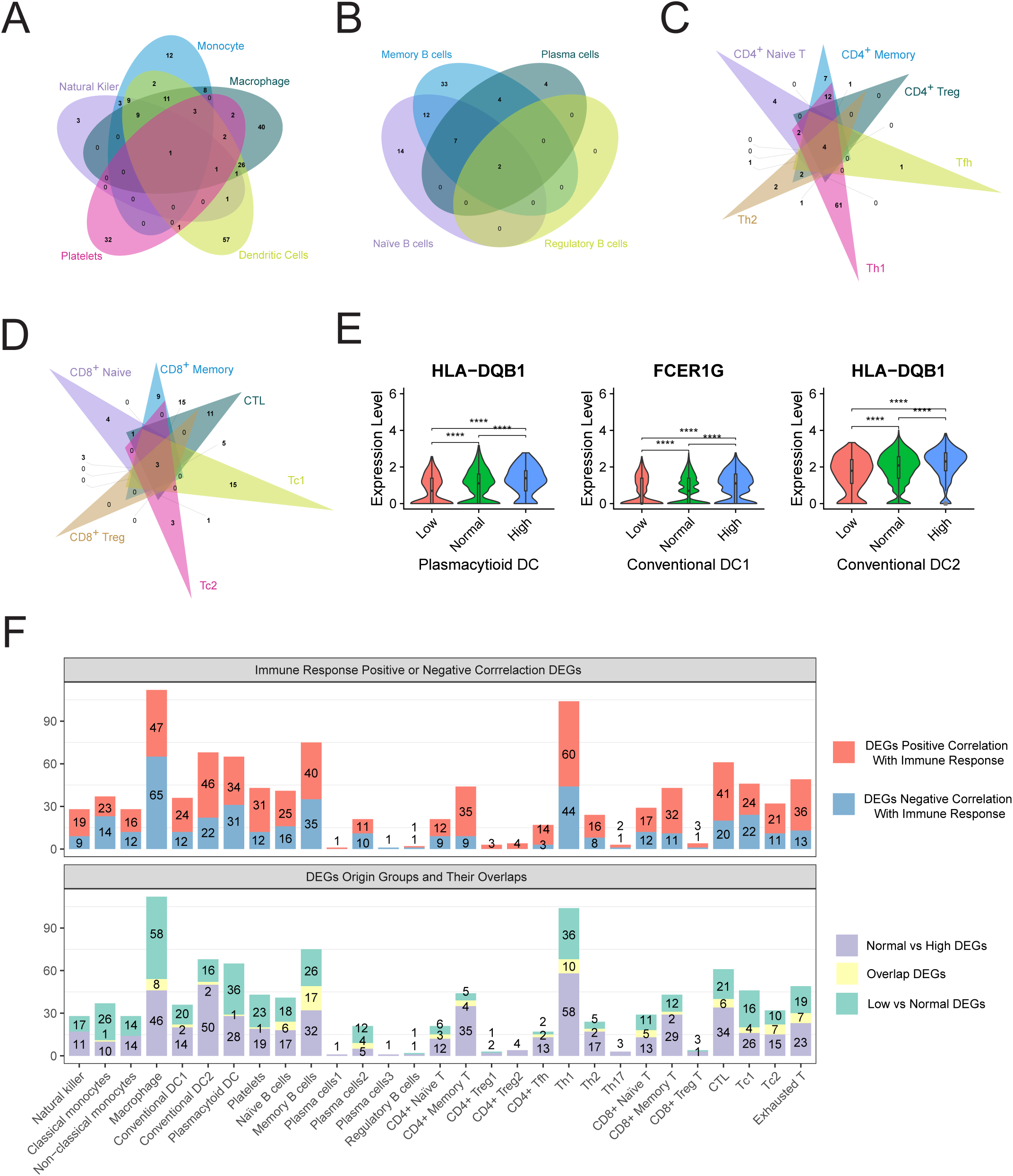
Differential Expression Genes (DEGs) Affecting Antibody Titers Across Various Cell Types. (A-D) Venn diagrams illustrating the DEGs that influence antibody titers. These diagrams are presented for collections of innate immune cells (A), B cells (B), CD4+ T cells (C), and CD8+ T cells (D). (E) Violin plot showed DEGs that exhibit significant differences across various antibody titer groups. (F) Displays the count of DEGs within different types of cells. The x-axis denotes the categories of cells, while the y-axis indicates the number of DEGs. In the upper panel, the counts of DEGs are distinguished by colors, with genes positively correlated with antibodies marked in red and those negatively correlated marked in blue. The lower panel uses different colors to indicate the origins of the DEGs identified. Genes originating from the Normal vs High group are highlighted in purple, those from the Low vs Normal group in green, and the intersection of these two groups is shown in yellow.

It’s easy to spot the traditional understanding of Th1 cells, memory B cells, and DC cells supports our finding of a relative important roles on antibody production post-vaccination: Th1 cells can enhance antibody production by stimulating B cell proliferation and differentiation[28]. Memory B cells rapidly produce antibodies with high affinity upon re-encountering antigens[29], while DC cells efficiently process and present antigens to activate T cells and B cells for antibody response and antibody production[30]. However, the high relative number of differentially expressed genes (DEGs) in macrophages is unexpected. Despite being a classic immune cell, there is little direct evidence linking macrophages to antibody production. Nevertheless, macrophages are considered the central players in the entire immune system, initiating and guiding almost all antibody responses, including those involving T cells and B cells/adaptive immunity.[31]. We will be comparing the differentially expressed gene (DEG) enriched pathways identified in macrophages with pathways enriched in other cell types later.

On the other hand, overall, there doesn’t seem to be a clear bias in the ratio of upregulated (positive correlation with antibody titer) and downregulated genes (negative correlation with antibody titer) during antibody response. Across different cell subtypes, the ratio generally does not exceed 1:3 or 3:1 (Fig2F). This indicates that antibody response regulation involves both upregulation and downregulation of genes across multiple cell types, rather than being solely influenced by one cell type. However, at the gene level, some genes show a strong association with antibody response across multiple cell types. For example, the HLA-DQB1 gene exhibits significant intergroup upregulation differences in both pDCs and cDCs (Fig2E). HLA-DQB1 encodes an MHC-II protein that presents exogenous antigens to CD4+ T cells, playing a crucial role in the immune system. Besides, previous studies have shown that HLA-DQB1 may influence humoral immune response elicited by inactivated Japanese encephalitis vaccine (IJEV) [32]. Genes of the MHC-II class are associated with high-frequency antibody responses to five out of nine recombinant proteins tested in Rondonia State, Brazil [33]. All of these findings indicate the potential role of HLA-DQB1 in influencing population-specific antibody variations following vaccination.

Finally, we identified shared and specific DEGs among different subtypes: MT-CO1 for innate immune cells, HLA-DQA2 and FOSB for B cells, TXNIP and RPS10-NUDT3 for CD4+ T cells, and MT-ND3, RPS10-NUDT3, and JUN for CD8+ T cells (F2A-E). These genes play roles in energy production, T cell functions, and adaptive immunity, indicating its potential associations with antibody production by regulation multiple cells.

### Results 3: Innate antibody response cells dominate the majority of pathways contributing to antibody expression variants among individual

Using gene annotation and enrichment analysis tool Metascape [34], we try to depict pathways influencing population-level antibody differences across multiple cell types. We utilized databases such as GO, KEGG, Reactome, WikiPathway, CORUM, and Canonical Pathways. In total, we obtained 614 enriched pathways (Table S4). To organize these pathways, Metascape employed hierarchical clustering based on pathway similarity, resulting in 20 pathway categories (Table S3). We classified these categories into three groups based on the proportion of enriched pathways in each cell type (innate immune cells, B cells, and T cells), as shown in Figure 3A. The first group consists of pathways enriched in almost all cell types, referred to as “Common Pathways.” The second group is primarily enriched in innate immune cells and B cells, with fewer pathways enriched in T cells. The third group is dominated by pathways enriched in innate immune cells. From the overall picture, we observed the involvement of monocytes, macrophages, and dendritic cells (DCs) in pathways across all 20 major terms (Fig3A). However, at the T cell level, there was a greater enrichment of pathways associated with cytotoxic T lymphocytes (CTLs) and Th1 cells. These findings suggest that innate antibody response cells, along with B cells, play a crucial role in driving the differences in antibody expression among populations, while T cells, particularly CTLs and Th1 cells, have specific pathway enrichments.

**Figure 3.**
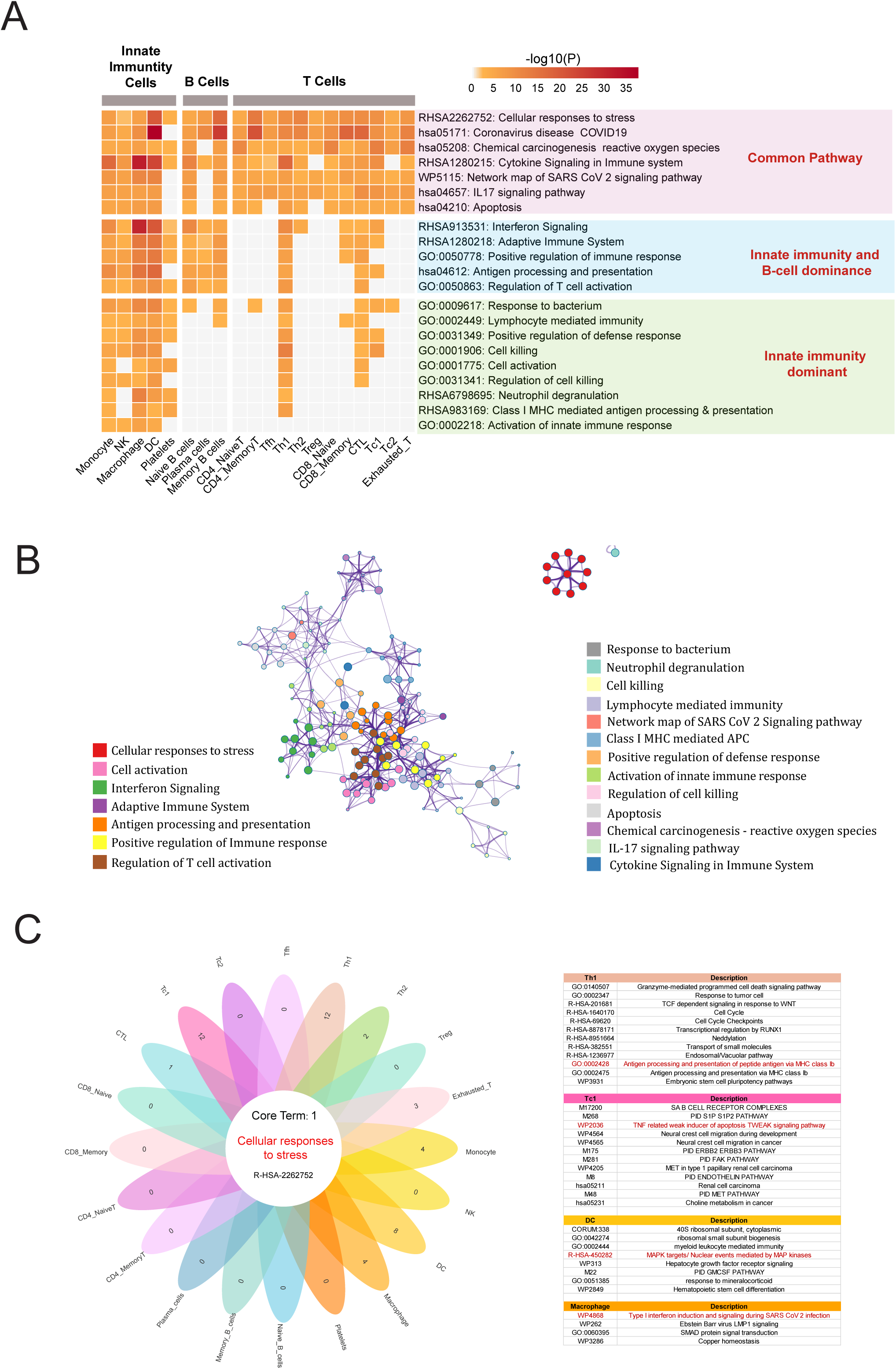
Enrichment Map of Differential Genes Influencing Antibody Expression in Various Cells. (A) Heatmap illustrating the enrichment of biological terms that influence antibody expression across different cell types. The x-axis lists the categories of cells, while the y-axis lists representative biological terms from similar term clusters. The color intensity within each cell of the heatmap signifies the level of enrichment significance, with gray indicating a lack of enrichment for that pathway in the specified cell type. (B) Network diagram that visualizes the enrichment biological terms impacting antibody expression in various cells. Pathways with similarities are grouped together. The size of each node within the network correlates with the degree of enrichment significance. Node colors are consistent with those in panel A, representing biological terms from different enrichment clusters. The number of nodes reflects the number of pathways associated with each term. (C) Petal diagram for the enrichment terms associated with biological terms that affect antibody expression in different cells. The center of the petal represents pathways shared across cell types, while the petals indicate the count of unique pathways specific to individual cell types. The right panel of this diagram showcases some significant cell-specific terms.

From the perspective of shared signaling pathways, the common pathway observed in all cells is the *cellular response to stress* (RHSA2262752). This term encompasses a complex biological process with diverse signal transduction pathways and molecular mechanisms, playing a regulatory role in both innate and adaptive immune responses [35]. Our finding indicates a potential link between the cellular stress response and antibody production post-vaccination, relevant to all peripheral blood cell types. Besides the cellular stress pathway, our findings also underscore the importance of effective cytokine regulation in developing a strong antibody response: Cytokine signaling emerges as a key shared pathway across various cells, highlighted in pathways like *Cytokine Signaling in the Immune System* (RHSA1280215), *Network Map of SARS-CoV-2 Signaling Pathway* (WP51150), and *IL-17 Signaling Pathway* (hsa04657) (Fig3A).

In the “Innate immune and B-cell dominated” pathway, pathways involving in this categorize primarily relate to interferon signaling and pathways associated with the adaptive immune system (such as T cell activation and antigen presentation) (Fig3A). In antibody responses. Interferons are mainly produced by innate immune cells, such as macrophages and monocytes[36]. Previous studies have shown that interferons play diverse roles in regulating B cell antibody production, including promoting B cell activation, enhancing antibody responses, increasing antibody production, regulating antibody class switching, enhancing antibody affinity, and balancing immune reactions[36–39]. Our results suggest that differential responses to interferon signaling in the population contribute to variations in antibody production among different individuals, primarily mediated by cells of the innate antibody response and B cells.

The last category is mainly composed of pathways related to innate immunity, including the *Activation of innate antibody responses* (GO: 000218), *Neutrophil degranulation* (RHSA6798695), *cell activation* (GO:0001775:), and *cell killing* (GO:0001906:). Our analysis results show that in these pathways, the non-innate immune cells typically include Th1, CTL, and Tc1. In terms of inter-pathway correlations, we observed that most pathways are strongly correlated (Fig3B). Antigen presentation, which is a crucial link connecting innate and adaptive immunity, is located in the center of the entire pathway regulatory network, which is not surprising. By presenting antigens to T cells, antigen presentation triggers subsequent adaptive antibody responses. Interestingly, the cellular stress response pathways are largely uncorrelated with the overall pathway network. This suggests that while cellular stress response pathways play important roles in maintaining cellular homeostasis and survival, they may primarily influence the inter-individual differences in antibody expression through mechanisms other than direct involvement in the core pathways of antibody responses.

In the immune system, different types of cells play unique roles and participate in antibody responses through specific signaling pathways. A total of eight cell types were identified, each enriched with cell type-specific pathways, including Th1 (12 pathways), Tc1 (12 pathways), DC (8 pathways), Monocyte (4 pathways), Macrophage (4 pathways), Exhausted T (3 pathways), Th2 (2 pathways), and CTL (1 pathway) (Fig3C). We then particularly focus in the specific enrichment pathway in macrophages, as they have the highest number of DEGs (differentially expressed genes) among cell types (Fig2F). We have discovered that macrophages exhibit a unique enrichment in *Type I interferon induction and signaling during SARS-CoV-2 infection* (WP4868). While previous research has linked the lack of the Type I interferon pathway to severe COVID-19 infections [40, 41], our findings suggest that the varying response of this pathway within macrophages in the population may account for differences observed following antibody vaccination.

### Result 4: Deciphering the Impact of Protein Complex Networks Produced by PBMC Cells on Antibody Expression Variability

After depicting the pathways enriched with genes that contribute to inter-individual differences in antibody expression, we further attempted to understand the interactions between the proteins encoded by these genes and their potential connection to antibody responses. Protein-protein interaction relationships were determined using metascape [34], which utilized the STRING and BioGrid databases [42, 43]. Additionally, the Molecular Complex Detection (MCODE) algorithm was employed to identify densely connected network components [44]. The analysis identified a total of eight MCODE protein complexes: MCODE1 ribosome complex network, MCODE2 interferon complex network, MCODE3 MHC complex network, MCODE4 inflammation response complex network, MCODE5 ATP complex network, MCODE6 RNA splicing complex network, MCODE7 B cell receptor complex network, and MCODE8 kinase transcription factor complex network (Fig4A).

**Figure 4.**
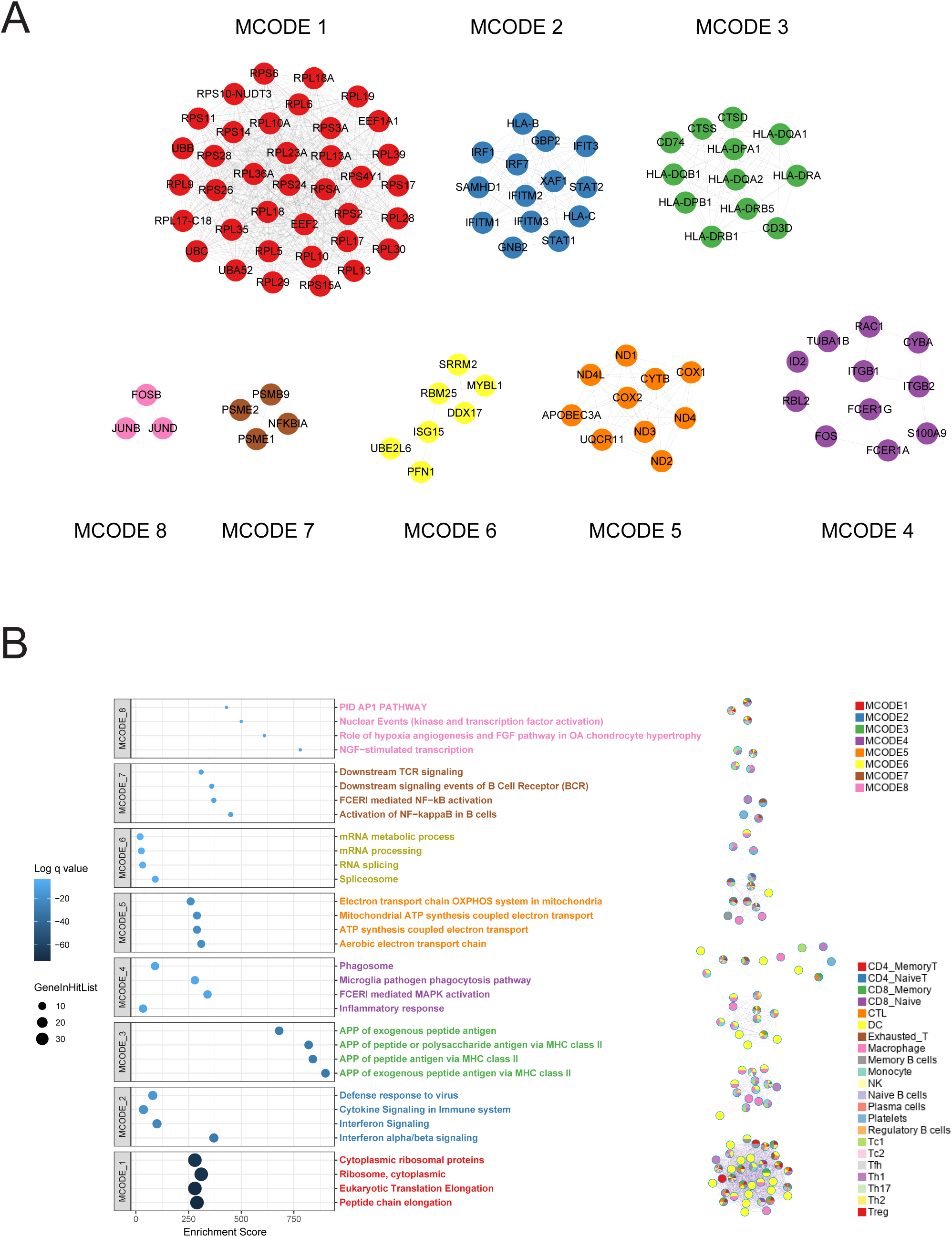
Protein Interaction Networks in Peripheral Blood Affecting Antibody Expression. (A) Protein complexes in peripheral blood affecting antibody expression predicted using the Molecular Complex Detection (MCODE) algorithm. Each complex is represented by a distinct color. Each node within these complexes corresponds to a protein that is translated from one of the DEGs previously identified on antibody expression. (B) The left panel presents a dot plot illustrating the biological pathways associated with the complexes identified in part A. The enrichment score is plotted on the x-axis, while various enrichment terms are listed on the y-axis. The size of each dot reflects the count of genes present in the corresponding enriched pathway, and the color of the dots indicates the significance level of enrichment. The right panel outlines the cellular origins of each protein within the complexes, with different cell types distinguished by unique colors.

Among these, the ribosome complex network involved the largest number of proteins and exhibited significantly higher enrichment compared to other complexes. By analyzing the cellular composition responsible for secreting these complexes, we found that although various cell types were involved, DC cells were notably the most prominently engaged (Fig4B). By targeting the MCODE1 ribosome complex network-related pathways, significant enrichment was observed in the *Eukaryotic Translation Elongation* (R-HSA-156842) and *peptide chain elongation* (R-HSA-156902) pathways (Fig4B). *Eukaryotic Translation Elongation* is a crucial phase in the synthesis of proteins, where *peptide chain elongation* involving the coordinated action of various ribosomal proteins and related factors. This ensures the accurate translation of mRNA-encoded amino acid sequences into corresponding proteins. This mechanism is vital for the normal functioning of all cells, including dendritic cells (DCs) within the immune system. DCs act as a pivotal link between innate and adaptive immunity; thus, any factor that affects DC functionality could significantly impact immune responses [45–47].

### Result5: Possible mechanism of Dendritic Cells on Antibody Production Through Ribosome Complex Network

Based on above findings, it’s plausible to suggest that dendritic cells (DCs), given their significant role in the ribosome complex network, could influence antibody production through various mechanisms:

1. Enhanced Antigen Presentation: DCs can boost cytokine secretion by TH1 cells and support the development and maturation of naive B cells. By controlling the production of specific proteins, DCs enhance their ability to process and present antigens more effectively. This leads to better activation of T cells and increased antibody production by B cells (Fig5). This mechanism serves as a key pathway by which DCs indirectly affect B cell antibody production [48, 49]. Our analysis also shows a higher proportion of complexes presented by MHCII are attributed to DCs (Fig4B), further supporting this pathway.
2. Cytokine Modulation: DCs can emit pro-inflammatory cytokines, such as IL-6 and IL-12 [50–52], which facilitate B cell class switching (Fig5). This action bolsters IgG secretion via the ribosome complex network.
3. Immune Memory Formation: DCs, through the ribosome complex network, contribute to the regulation of immune memory cell generation [53]. This is crucial for sustaining long-term antibody responses. While antibodies have a finite lifespan, memory B cells can persistently activate to produce new plasma cells, keeping antibody levels against specific pathogens steady. Our data indicate that samples with high antibody expression 21 days post-second vaccination maintained elevated antibody levels at 4 months. However, it’s important to note that this process predominantly occurs in the germinal centers (GC) of lymph nodes and not in the peripheral blood, hence it’s not depicted in Fig5.
4. Direct B Cell Maturation Induction: Evidence suggests that DCs can directly induce B cell maturation through receptor CD40 [54], including instances where conventional DCs (cDCs) present antigens directly to B cells [55].

**Figure 5:**
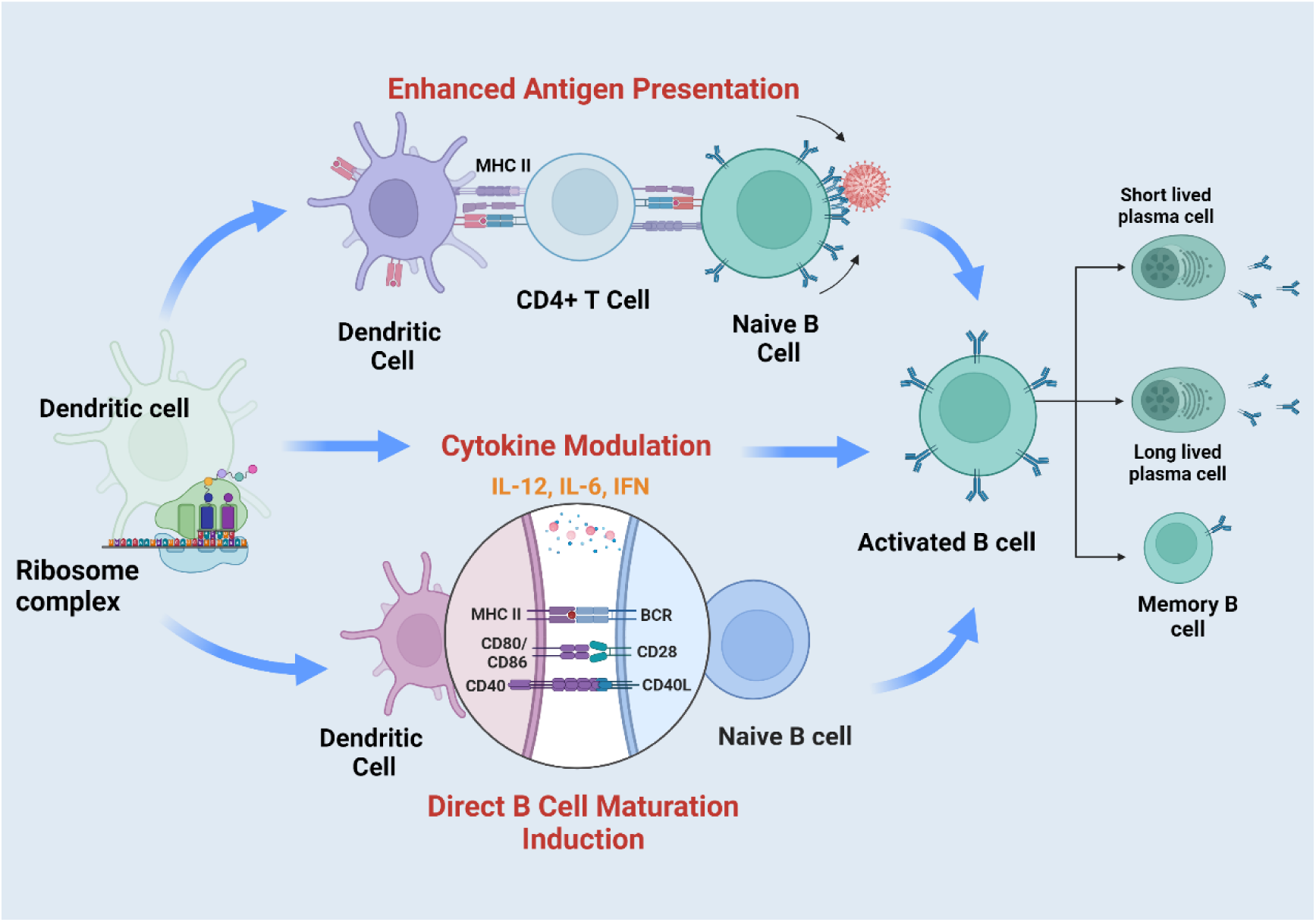
Dendritic Cells (DCs) and Their Role in Ribosome Complex Network-Mediated Antibody Production. This figure showcases the complex influence of dendritic cells (DCs) within the ribosome complex network on antibody secretion. It highlights three primary mechanisms through which DCs can either directly or indirectly affect B cell antibody production: 1). Enhanced Antigen Presentation: Illustration includes a DC presenting antigens to a T helper cell (TH1) via MHC II. This process fosters interactions between activated T cells and naive B cells, ultimately leading to B cell maturation and an increase in antibody production. 2). Cytokine Modulation: Depicts a DC releasing pro-inflammatory cytokines (IL-6 and IL-12), which facilitate B cell class switching and the secretion of IgG antibodies. 3). Direct B Cell Maturation Induction: Shows a DC directly engaging with a B cell through the CD40 receptor, CD80-CD28 interaction, or by presenting antigens to B cells, thereby inducing B cell maturation. Each pathway underscores the significant regulatory capabilities of DCs in modulating immune responses and enhancing antibody secretion. Figure Created with BioRender.com, with permission.

In summary, our research demonstrates that the ribosome complex network, primarily facilitated by DCs, plays a crucial role in regulating antibody production. This underscores the intricate nature of cellular interactions and the critical need for precise regulation within the immune system.

## Discussion

The study of differences in antibody production among different populations after vaccination is a complex issue that involves multiple dimensions such as genetic background, environmental factors, and lifestyle. With the rapid advancement of biotechnology, single-cell transcriptomics has emerged as a powerful tool for gaining a deeper understanding of the fundamental reasons behind these differences. By measuring RNA expression at the single-cell level, single-cell transcriptomics provides a unique approach to analyze the functions and states of cells, revealing unique expression patterns of different cell types in complex biological tissues and how they respond under various physiological and pathological conditions.

In the exploration of antibody generation following vaccination, the application of single-cell transcriptomics has demonstrated its unique value. Firstly, it enables the identification of key cell subtypes involved in the antibody response, including rare cell types that may be overlooked in population-level analyses. This detailed analysis provides a more comprehensive understanding of the antibody response. Secondly, by comparing the gene expression differences of the same cell types among different individuals, single-cell transcriptomics helps identify key genes and signaling pathways that may influence antibody production. This is crucial for uncovering the molecular mechanisms underlying differences in antibody production. Finally, by observing changes in cellular transcriptomes after vaccination, single-cell transcriptomics can help us gain a deeper understanding of the regulatory mechanisms of the antibody response. This includes identifying the cell types and molecules involved in antibody production and regulation, thus providing valuable insights into how the immune system responds to vaccination.

Analyzing transcriptome data four months after vaccination provides a unique perspective in understanding the differences in antibody generation among different populations. This time frame allows the immune system to mount an initial response to the vaccine and enter a relatively stable memory phase, reflecting the variation in immune memory formation and antibody production among individuals. By analyzing the transcriptome at this crucial time point, researchers can deeply evaluate the formation of long-term immune memory. This includes observing the transcriptomic changes in memory cells and other key immune cells to understand how immune memory is established in different populations, revealing factors that influence long-term immune protection. Additionally, this analysis can help identify factors that continue to impact antibody levels post-vaccination, such as persistent gene expression patterns and cellular state changes. This may include chronic inflammatory responses or alterations in cellular metabolism, both of which could have significant effects on antibody production and persistence. Furthermore, through transcriptome analysis, researchers can track potential mechanisms leading to immune evasion. If differences in antibody production exist, this analysis can uncover potential reasons for these differences, such as changes in antigen-presenting cell function, which are crucial for understanding individual variations in vaccine efficacy.

This study employs single-cell transcriptomic sequencing technology to conduct innovative research aimed at revealing the molecular mechanisms underlying differences in antibody production following vaccination. The main findings of this study cover several key aspects. Firstly, the research successfully identified genes and cell subpopulations closely associated with differences in antibody production among individuals. This discovery highlights the roles of specific genes and cell subpopulations in the antibody generation process and emphasizes the importance of innate antibody response cells, providing a new perspective on understanding differences in antibody responses among different populations. Secondly, the study identified the central role of the ribosomal complex network in regulating antibody production. Through analysis of peripheral blood transcriptomic expression complexes, the research revealed the potential key role played by the ribosomal complex network, predominantly led by dendritic cells (DCs), in regulating antibody production. This finding presents new targets for future design of targeted immune intervention strategies, particularly in optimizing vaccine design (such as considering the targeted addition or enhancement of certain cell types or pathways to achieve better antibody response effects, and in improving immune efficiency) and enhancing immune efficiency.

Of particular note is the significant potential of single-cell transcriptomic data in evaluating cytokine profiles and assessing immune storm occurrences post-vaccination. Cytokines, as messengers in the immune system, reflect the immune system’s active state and potential abnormal reactions through changes in their expression patterns. Through single-cell transcriptomic analysis, we can precisely identify the activated cell subpopulations responding post-vaccination and the specific cytokines they secrete. This not only aids in a better understanding of how vaccines trigger protective antibody responses but also helps us predict and mitigate potential adverse reactions, such as immune storms. Furthermore, by integrating single-cell transcriptomic data with traditional antibody response assessment data, we can construct more comprehensive and precise antibody response models. These models not only provide crucial biological foundations for vaccine design and assessment but also offer guidance for future personalized immunotherapy strategies.

In conclusion, while directly predicting specific antibody levels using single-cell transcriptomic data currently faces technical and data challenges, our research demonstrates the vast potential of single-cell transcriptomics in deeply understanding the mechanisms of antibody responses triggered by vaccines. We believe that with technological advancements and the accumulation of more high-quality data, single-cell transcriptomics will play an increasingly important role in future vaccine research and immunotherapy fields.

## Method

### Study subjects

Healthy adult individuals who had received the COVID-19 vaccine were recruited as study subjects. In order to ensure data reliability and consistency, the following inclusion criteria were established: 1) participants with no prior SARS-CoV-2 infection who had completed the two-dose regimen of the inactivated COVID-19 vaccine; 2) the interval between the initial and booster doses was 3 to 4 weeks; 3) the final vaccination occurred between July 15, 2020, and August 30, 2020. Details including vaccination dates, gender, and age were recorded for each participant, with blood samples collected 21 days post-second dose and between 135 to 140 days post-vaccination to assess serum IgG antibody levels. Blood collection took place at BGI YouKang Clinic. All participants provided informed consent, and the study was approved by the BGI Institutional Review Board (approval number BGI-IRB 20161).

### Antibody Detection

The SARS-CoV-2 RBD/S antibody detection kit (HWTS-RT055A, Macro & Micro-Test) was used to identify SARS-COV-2 specific IgG antibodies. This kit operates on a double-antigen sandwich enzyme-linked immunosorbent assay principle, with the SARS-CoV-2 RBD antigen initially coated on the microplate. Following this, blood samples are diluted in gradients and added to the microplate, then incubated with an enzyme-labeled antigen. In the presence of antibodies against the SARS-CoV-2 S protein RBD region, a “coated RBD antigen-SARS-CoV-2 antibody-enzyme-labeled RBD antigen” complex is formed. Addition of a chromogenic substrate leads to the enzyme catalyzing the production of a blue substrate. The reaction is halted, resulting in the substrate turning yellow. If RBD or S antibodies are absent in the sample, no color change occurs. The ELISA reader measures the OD value of the reaction, and positivity or negativity is determined based on a predefined cutoff value. The reciprocal of the dilution at which the OD falls below the critical reference value corresponds to the antibody titer in the serum.

### Peripheral blood mononuclear cell (PBMC) collection

For each participate, Peripheral blood samples (3 mL) were collected into EDTA anticoagulant tubes and gently mixed by inverting 4-6 times. The whole blood was then diluted with 3 mL of phosphate-buffered saline (PBS) and transferred to a 15 mL centrifuge tube. Using Ficoll-Paque Plus (Sigma Aldrich) solution, PBMCs were isolated following standard density gradient centrifugation methods. Cell collection and counting were performed using the Cellaca MX high-throughput cell counter (Nexcelom Bioscience). The isolated PBMCs were resuspended in a freezing medium consisting of 90% fetal bovine serum (FBS, HyClone) and 10% DMSO, followed by freezing at −80°C for 24 hours using Nalgene® Mr. Frosty Cryo 1°C freezing containers (Thermo Fisher Scientific) before long-term storage in liquid nitrogen. All procedures were carried out under sterile conditions.

### Single-nucleus suspension preparation

Frozen PBMC storage vials were rapidly thawed in a 37°C water bath for approximately 2 minutes until only a small ice crystal remained. Thawed PBMCs were quenched with 4 mL of pre-warmed 1X phosphate-buffered saline (PBS, Thermo Fisher Scientific) at 37°C and supplemented with 10% FBS. The mixture was centrifuged at 500 x g for 10 minutes at room temperature. The supernatant was removed, and the cell pellet was resuspended in 3 mL of 1X PBS containing 0.04% bovine serum albumin (BSA, Sangon Biotech).

The resuspended cells were filtered through a 40 μm cell strainer (Falcon) and then subjected to centrifugation. Following the manufacturer’s protocol, dead cells were removed using magnetic bead purification (Miltenyi Biotech) before proceeding to single-cell RNA sequencing (scRNA-seq). The cells were resuspended in a cell resuspension buffer to achieve a concentration of 1000 cells/μL.

### Library construction and Single-cell RNA-seq

We prepared scRNA-seq libraries using the DNBelab C single-cell library preparation kit (MGI, #1000021082) following previously reported methods. Briefly, cell nuclei were resuspended in a PBS solution containing 0.04% BSA and filtered through a 40μm cell strainer. The concentration of the cell suspension was recorded and measured for the use of DNBelab C single-cell library preparation kit (MGI, #1000021082) to generate droplets. After collecting the mixed droplets, emulsion breaking was performed, followed by reverse transcription, cDNA and oligonucleotide amplification, and product filtration. Oligonucleotide products were then barcode-labeled for PCR to create oligonucleotide circular libraries, while cDNA products were prepared into single-stranded DNA libraries. Finally, sequencing was conducted at the China National GeneBank (Shenzhen) using the DNBSEQ-T1 sequencer with a read length of 30bp for reads1 and 100bp for reads2.

### Single-cell RNA-seq data processing

The raw sequencing reads of DIPSEQ-T1 were filtered and demultiplexed by PISA (v.0.2) (https://github.com/shiquan/PISA). The reads were aligned to the human genome (GRCh38_release95) using STAR (v.2.7.4a)[56] and sorted using Sambamba (v.0.7.0)[57]. Doublet and contaminant cells were rigorously filtered based on clustering in the UMAP embedding space, unmarked expression, and estimation using DoubletFinder[58]. Cells with feature counts below 200 or above 4000 (filtered out) and cells with mitochondrial UMI counts exceeding 10% (filtered out) were removed from the gene-cell matrix. Prior to filtering, the median gene count per cell across all cells was 3628 [IQR 2118-5497], and the median feature count was 1222 [IQR 848-1625]. After filtering, the median gene count per cell across all cells was 3768 [IQR 2287–5600], and the median feature count was 1253 [IQR 890–1645]. The SCTransform function in Seurat (version 4.1.1) was performed to normalize the data for further clustering purposes.

### Cell clustering, annotation, and DEGs identify

PCA and UMAP were used for dimensionality reduction in Seurat (version 4.1.1)[59]. Based on markerset1 in the Supplementally Table 1, the first 20 principal components were utilized for UMAP projection and clustering analysis to differentiate innate immune cells, B cells, and T cells into three clusters. Subsequently, the Innate cells subtype marker set (dim=10, resolution=0.2), B cells subtype marker set (dim=15, resolution=0.2), CD4+ T cell subtype marker set (dim=14, resolution=0.4), and CD8+ T cell subtype marker set (dim=10, resolution=0.4) were used to identify subtypes within the three clusters. Following subtype identification, the barcode information from each cell-id was used to mapped back to the original three-cluster plot to visualize the cellular subtypes. For differential gene identification, the Seurat FindAllMarkers function was employed to recognize differentially expressed genes between clusters, with the parameters set as pos = TRUE, min.pct = 0.25, logfc.threshold = 0.25, test.use = ’wilcox’. Genes with an adjusted p-value < 0.01 were considered differentially expressed.

For achieve the goal of annotation of classical PBMC cell types, we employed a three-step approach. Firstly, we used a set of markers to classify all cells into innate immune cells, B cells, and T cells (Figure S6). Secondly, we further subdivided these three major cell types into subtypes by extracting different clusters and using distinct subtype markers (Figures 1C-F and Figure S8-S15). After subtyping, in the third step, we remapped back to the original clustering diagram for visualization, resulting in Figure 1B. It is important to note that the results of differential gene expression and cell composition analysis presented in this study are based on cell subtype classification and not on the clustering visualization in Figure 1B. We believe that the results obtained by further subclassifying cell types using subtype markers, although eliminating cells that cannot be differentiated using subtype markers, are more accurate due to the use of additional subtype markers and finer subdivision within subtypes. Moreover, in subtype annotation, we also considered the issue of cell type proportions, enhancing the accuracy of subtype analysis. Specific code can be found in the Code Available Part.

### Differential Gene Enrichment and Protein Interaction Analysis

The enrichment analysis of the differentially expressed genes in the transcriptome was conducted using the Metascape software[34]. Specifically, the differential genes from various cell types were tabulated and uploaded to the Metascape online software, and the resulting data was further processed using default parameters. For the differential genes in different cell types, we utilized Venn diagrams, violin plots, and bar charts for visualization. The Venn diagram was created using the online software Eveen[60], the violin plot used the built-in function VlinPlot in Seurat, and the bar chart was generated using ggplot2[61].

By calculating the proportion of enriched pathways across all cell types, the representative 20 pathways were categorized into three groups: Common Pathway, Innate Immunity and B-cell Dominance, and Innate Immunity Dominant. The selection of representative pathways was based on the significance of the p-value and the cell type proportion. The pathway enrichment heatmap was plotted using the pheatmap (version 1.0.12) package in R. The network enrichment diagram was directly produced by Metascape, while the petal diagram was drawn using Eveen.

The MCODE network diagram was created by downloading the network generated by Metascape and visualizing it in Cytoscape[62]. The bubble charts for the pathways involved in each MCODE were plotted using R package ggplot2[61].

## Declarations

### Ethics approval and consent to participate

This study was approved by the BGI Institutional Review Board with approval number BGI-IRB 20161. Written informed consent for the storage, detection and potential research usage of the blood samples for further studies was obtained from the participants before enrollment.

### Availability of data and materials

Single cell transcriptome data used in this project are deposited to the CNGB Sequence Archive (CNSA) of China National GeneBank DataBase (CNGBdb) with accession number CNP0005357. (https://db.cngb.org/search/project/CNP0005357/)

### Author contributions

Conceptualization, Jinmin Ma, Weijun Chen and Zhongyi Zhu; Data curation, Jiatong Sun, Yong Chen and Lei Zhang; Formal analysis, Zhongyi Zhu, Jiatong Sun, Lei Zhang and Fubaoqian Huang; Funding acquisition, Jinmin Ma; Investigation, Zhongyi Zhu and Meirong Li; Methodology, Yaling Huang; Project administration, Jinmin Ma; Software, Fubaoqian Huang; Supervision, Chuanyu Liu and Weijun Chen; Validation, Yaling Huang; Writing – original draft, Zhongyi Zhu; Writing – review & editing, Meirong Li and Jinmin Ma.

### Code Available

All code used to replicate the results of the study analysis can be found on GitHub Repositories under the Apache License 2.0: https://github.com/zzy221127/SingleCellVaccine_pub

## Acknowledgement

We express our gratitude to BGI YouKang Clinic and BGI Clinical Laboratories (Shenzhen) Co., Ltd. for their efforts in collecting blood samples, sample information, and conducting antibody detection.

## Reference

[1] K. A. Earle et al., “Evidence for antibody as a protective correlate for COVID-19 vaccines,” Vaccine, vol. 39, no. 32, pp. 4423–4428, Jul 22 2021.

[2] D. S. Khoury et al., “Neutralizing antibody levels are highly predictive of immune protection from symptomatic SARS-CoV-2 infection,” Nat Med, vol. 27, no. 7, pp. 1205-1211, Jul 2021.

[3] P. B. Gilbert et al., “Immune correlates analysis of the mRNA-1273 COVID-19 vaccine efficacy clinical trial,” Science, vol. 375, no. 6576, pp. 43–50, Jan 7 2022.

[4] D. S. Khoury et al., “Correlates of Protection, Thresholds of Protection, and Immunobridging among Persons with SARS-CoV-2 Infection,” Emerg Infect Dis, vol. 29, no. 2, pp. 381-388, Feb 2023.

[5] U. Wiedermann, E. Garner-Spitzer, and A. Wagner, “Primary vaccine failure to routine vaccines: Why and what to do?,” Hum Vaccin Immunother, vol. 12, no. 1, pp. 239–43, 2016.

[6] J. L. Lee and M. A. Linterman, “Mechanisms underpinning poor antibody responses to vaccines in ageing,” Immunol Lett, vol. 241, pp. 1-14, Jan 2022.

[7] P. Naaber, et al., “Dynamics of antibody response to BNT162b2 vaccine after six months: a longitudinal prospective study,” Lancet Reg Health Eur, vol. 10, p. 100208, Nov 2021.

[8] C. Munro, “Covid-19: 40% of patients with weakened immune system mount lower response to vaccines,” BMJ, vol. 374, p. n2098, Aug 24 2021.

[9] L. M. Frommert et al., “Type of vaccine and immunosuppressive therapy but not diagnosis critically influence antibody response after COVID-19 vaccination in patients with rheumatic disease,” RMD Open, vol. 8, no. 2, Dec 2022.

[10] H. Amellal, N. Assaid, K. Akarid, A. Maaroufi, S. Ezzikouri, and M. Sarih, “Mix-and-match COVID-19 vaccines trigger high antibody response after the third dose vaccine in Moroccan health care workers,” Vaccine X, vol. 14, p. 100288, Aug 2023.

[11] F. A. Pearce, et al., “Antibody prevalence after three or more COVID-19 vaccine doses in individuals who are immunosuppressed in the UK: a cross-sectional study from MELODY,” Lancet Rheumatol, vol. 5, no. 8, pp. e461–e473, Aug 2023.

[12] B. Pulendran, S. Li, and H. I. Nakaya, “Systems vaccinology,” Immunity, vol. 33, no. 4, pp. 516–29, Oct 29 2010.

[13] D. A. Collier et al., “Age-related immune response heterogeneity to SARS-CoV-2 vaccine BNT162b2,” Nature, vol. 596, no. 7872, pp. 417–422, Aug 2021.

[14] A. Lee et al., “Efficacy of covid-19 vaccines in immunocompromised patients: systematic review and meta-analysis,” BMJ, vol. 376, p. e068632, Mar 2 2022.

[15] C. A. Martin, et al., “Ethnic differences in cellular and humoral immune responses to SARS-CoV-2 vaccination in UK healthcare workers: a cross-sectional analysis,” EClinicalMedicine, vol. 58, p. 101926, Apr 2023.

[16] O. V. Adeniyi, O. C. Durojaiye, and C. Masilela, “Persistence of SARS-CoV-2 IgG Antibody Response among South African Adults: A Prospective Cohort Study,” Vaccines (Basel), vol. 11, no. 6, Jun 6 2023.

[17] D. D. Chaplin, “Overview of the immune response,” J Allergy Clin Immunol, vol. 125, no. 2 Suppl 2, pp. S3-23, Feb 2010.

[18] R. Carsetti et al., “Comprehensive phenotyping of human peripheral blood B lymphocytes in healthy conditions,” Cytometry A, vol. 101, no. 2, pp. 131-139, Feb 2022.

[19] B. Hwang, J. H. Lee, and D. Bang, “Single-cell RNA sequencing technologies and bioinformatics pipelines,” Exp Mol Med, vol. 50, no. 8, pp. 1-14, Aug 7 2018.

[20] D. Jovic, X. Liang, H. Zeng, L. Lin, F. Xu, and Y. Luo, “Single-cell RNA sequencing technologies and applications: A brief overview,” Clin Transl Med, vol. 12, no. 3, p. e694, Mar 2022.

[21] D. Lahnemann et al., “Eleven grand challenges in single-cell data science,” Genome Biol, vol. 21, no. 1, p. 31, Feb 7 2020.

[22] A. Adil, V. Kumar, A. T. Jan, and M. Asger, “Single-Cell Transcriptomics: Current Methods and Challenges in Data Acquisition and Analysis,” Front Neurosci, vol. 15, p. 591122, 2021.

[23] S. A. CZI Single-Cell Biology Program, Brian Aevermann, Pedro Assis, Seve Badajoz, Sidney M. Bell, Emanuele Bezzi, Batuhan Cakir, Jim Chaffer, Signe Chambers, J. Michael Cherry, Tiffany Chi, Jennifer Chien, Leah Dorman, Pablo Garcia-Nieto, Nayib Gloria, Mim Hastie, Daniel Hegeman, Jason Hilton, Timmy Huang, Amanda Infeld, Ana-Maria Istrate, Ivana Jelic, Kuni Katsuya, Yang Joon Kim, Karen Liang, Mike Lin, Maximilian Lombardo, Bailey Marshall, Bruce Martin, Fran McDade, Colin Megill, Nikhil Patel, Alexander Predeus, Brian Raymor, Behnam Robatmili, Dave Rogers, Erica Rutherford, Dana Sadgat, Andrew Shin, Corinn Small, Trent Smith, Prathap Sridharan, Alexander Tarashansky, Norbert Tavares, Harley Thomas, Andrew Tolopko, Meghan Urisko, Joyce Yan, Garabet Yeretssian, Jennifer Zamanian, Arathi Mani, Jonah Cool, Ambrose Carr, “CZ CELL×GENE Discover: A single-cell data platform for scalable exploration, analysis and modeling of aggregated data,” bioRxiv, 2023-01-01 00:00:00 2023.

[24] C. Dominguez Conde et al., “Cross-tissue immune cell analysis reveals tissue-specific features in humans,” Science, vol. 376, no. 6594, p. eabl5197, May 13 2022.

[25] S. C. De Rosa, “Vaccine applications of flow cytometry,” Methods, vol. 57, no. 3, pp. 383–91, Jul 2012.

[26] E. Gianchecchi, A. Torelli, P. Piu, C. Bonifazi, L. Ganfini, and E. Montomoli, “Flow cytometry as an integrative method for the evaluation of vaccine immunogenicity: A validation approach,” Biochem Biophys Rep, vol. 34, p. 101472, Jul 2023.

[27] R. Ran and D. K. Brubaker, “Enhanced annotation of CD45RA to distinguish T cell subsets in single-cell RNA-seq via machine learning,” Bioinform Adv, vol. 3, no. 1, p. vbad159, 2023.

[28] A. G. Telser, J Shock, “Molecular Biology of the Cell, 4th Edition,” vol. 18, p. 289, 2002.

[29] F. W. Alt, 本. 佑, A. Radbruch, and M. Reth, “Molecular biology of B cells,” 2015.

[30] W. R. Heath, Y. Kato, T. M. Steiner, and I. Caminschi, “Antigen presentation by dendritic cells for B cell activation,” Curr Opin Immunol, vol. 58, pp. 44–52, Jun 2019.

[31] C. D. Mills, “Anatomy of a discovery: m1 and m2 macrophages,” Front Immunol, vol. 6, p. 212, 2015.

[32] Y. Yao et al., “HLA Class II Genes HLA-DRB1, HLA-DPB1, and HLA-DQB1 Are Associated With the Antibody Response to Inactivated Japanese Encephalitis Vaccine,” Front Immunol, vol. 10, p. 428, 2019.

[33] J. C. Lima-Junior et al., “Influence of HLA-DRB1 and HLA-DQB1 alleles on IgG antibody response to the P. vivax MSP-1, MSP-3alpha and MSP-9 in individuals from Brazilian endemic area,” PLoS One, vol. 7, no. 5, p. e36419, 2012.

[34] Y. Zhou et al., “Metascape provides a biologist-oriented resource for the analysis of systems-level datasets,” Nat Commun, vol. 10, no. 1, p. 1523, Apr 3 2019.

[35] S. Muralidharan and P. Mandrekar, “Cellular stress response and innate immune signaling: integrating pathways in host defense and inflammation,” J Leukoc Biol, vol. 94, no. 6, pp. 1167–84, Dec 2013.

[36] F. McNab, K. Mayer-Barber, A. Sher, A. Wack, and A. O’Garra, “Type I interferons in infectious disease,” Nat Rev Immunol, vol. 15, no. 2, pp. 87–103, Feb 2015.

[37] C. L. Swanson, T. J. Wilson, P. Strauch, M. Colonna, R. Pelanda, and R. M. Torres, “Type I IFN enhances follicular B cell contribution to the T cell-independent antibody response,” J Exp Med, vol. 207, no. 7, pp. 1485-500, Jul 5 2010.

[38] M. Syedbasha, F. Bonfiglio, J. Linnik, C. Stuehler, D. Wuthrich, and A. Egli, “Interferon-lambda Enhances the Differentiation of Naive B Cells into Plasmablasts via the mTORC1 Pathway,” Cell Rep, vol. 33, no. 1, p. 108211, Oct 6 2020.

[39] A. Le Bon, G. Schiavoni, G. D’Agostino, I. Gresser, F. Belardelli, and D. F. Tough, “Type i interferons potently enhance humoral immunity and can promote isotype switching by stimulating dendritic cells in vivo,” Immunity, vol. 14, no. 4, pp. 461–70, Apr 2001.

[40] P. Bastard et al., “Autoantibodies against type I IFNs in patients with life-threatening COVID-19,” Science, vol. 370, no. 6515, Oct 23 2020.

[41] Y. M. Kim and E. C. Shin, “Type I and III interferon responses in SARS-CoV-2 infection,” Exp Mol Med, vol. 53, no. 5, pp. 750–760, May 2021.

[42] D. Szklarczyk, et al., “The STRING database in 2023: protein-protein association networks and functional enrichment analyses for any sequenced genome of interest,” Nucleic Acids Res, vol. 51, no. D1, pp. D638–D646, Jan 6 2023.

[43] R. Oughtred et al., “The BioGRID database: A comprehensive biomedical resource of curated protein, genetic, and chemical interactions,” Protein Sci, vol. 30, no. 1, pp. 187–200, Jan 2021.

[44] G. D. Bader and C. W. Hogue, “An automated method for finding molecular complexes in large protein interaction networks,” BMC Bioinformatics, vol. 4, p. 2, Jan 13 2003.

[45] X. Zhou, W. J. Liao, J. M. Liao, P. Liao, and H. Lu, “Ribosomal proteins: functions beyond the ribosome,” J Mol Cell Biol, vol. 7, no. 2, pp. 92–104, Apr 2015.

[46] L. Jiao, et al., “Ribosome biogenesis in disease: new players and therapeutic targets,” Signal Transduct Target Ther, vol. 8, no. 1, p. 15, Jan 9 2023.

[47] C. Petibon, M. Malik Ghulam, M. Catala, and S. Abou Elela, “Regulation of ribosomal protein genes: An ordered anarchy,” Wiley Interdiscip Rev RNA, vol. 12, no. 3, p. e1632, May 2021.

[48] D. Y. Tesfaye, A. Gudjonsson, B. Bogen, and E. Fossum, “Targeting Conventional Dendritic Cells to Fine-Tune Antibody Responses,” Front Immunol, vol. 10, p. 1529, 2019.

[49] S. B. Boscardin et al., “Antigen targeting to dendritic cells elicits long-lived T cell help for antibody responses,” J Exp Med, vol. 203, no. 3, pp. 599-606, Mar 20 2006.

[50] Y. D. Xu, M. Cheng, P. P. Shang, and Y. Q. Yang, “Role of IL-6 in dendritic cell functions,” J Leukoc Biol, vol. 111, no. 3, pp. 695-709, Mar 2022.

[51] B. Dubois et al., “Critical role of IL-12 in dendritic cell-induced differentiation of naive B lymphocytes,” J Immunol, vol. 161, no. 5, pp. 2223-31, Sep 1 1998.

[52] G. Jego, V. Pascual, A. K. Palucka, and J. Banchereau, “Dendritic cells control B cell growth and differentiation,” Curr Dir Autoimmun, vol. 8, pp. 124–39, 2005.

[53] M. Akkaya, K. Kwak, and S. K. Pierce, “B cell memory: building two walls of protection against pathogens,” Nat Rev Immunol, vol. 20, no. 4, pp. 229–238, Apr 2020.

[54] E. A. Clark, “Regulation of B lymphocytes by dendritic cells,” J Exp Med, vol. 185, no. 5, pp. 801-3, Mar 3 1997.

[55] T. M. Steiner, W. R. Heath, and I. Caminschi, “The unexpected contribution of conventional type 1 dendritic cells in driving antibody responses,” Eur J Immunol, vol. 52, no. 2, pp. 189–196, Feb 2022.

[56] A. Dobin, et al., “STAR: ultrafast universal RNA-seq aligner,” Bioinformatics, vol. 29, no. 1, pp. 15–21, Jan 1 2013.

[57] A. Tarasov, A. J. Vilella, E. Cuppen, I. J. Nijman, and P. Prins, “Sambamba: fast processing of NGS alignment formats,” Bioinformatics, vol. 31, no. 12, pp. 2032-4, Jun 15 2015.

[58] C. S. McGinnis, L. M. Murrow, and Z. J. Gartner, “DoubletFinder: Doublet Detection in Single-Cell RNA Sequencing Data Using Artificial Nearest Neighbors,” Cell Syst, vol. 8, no. 4, pp. 329–337 e4, Apr 24 2019.

[59] Y. Hao et al., “Integrated analysis of multimodal single-cell data,” Cell, vol. 184, no. 13, pp. 3573–3587 e29, Jun 24 2021.

[60] T. Chen, H. Zhang, Y. Liu, Y. X. Liu, and L. Huang, “EVenn: Easy to create repeatable and editable Venn diagrams and Venn networks online,” J Genet Genomics, vol. 48, no. 9, pp. 863–866, Sep 20 2021.

[61] P. M. Valero-Mora, “ggplot2: Elegant Graphics for Data Analysis,” Journal of Statistical Software, Book Reviews, vol. 35, no. 1, pp. 1–3, 07/30 2010.

[62] P. Shannon, et al., “Cytoscape: a software environment for integrated models of biomolecular interaction networks,” Genome Res, vol. 13, no. 11, pp. 2498–504, Nov 2003.

